# Genomic analysis of Strongyloides stercoralis and Strongyloides fuelleborni in Bangladesh

**DOI:** 10.1101/2024.05.16.24307305

**Authors:** Veroni de Ree, Tilak Chandra Nath, Dorothee Harbecke, Dongmin Lee, Christian Rödelsperger, Adrian Streit

## Abstract

**Background:** About 600 million people are estimated to be infected with *Strongyloides stercoralis*, the species that causes the vast majority of human strongyloidiasis cases. *S. stercoralis* can also infect non-human primates (NHPs), dogs and cats, rendering these animals putative sources for zoonotic human *S. stercoralis* infection. *S. fuelleborni* is normally found in old world NHPs but occasionally also infects humans, mainly in Africa. Dogs in southeast Asia carry at least two types of *Strongyloides*, only one of which appears to be shared with humans (“dog only” and “human and dog” types). For *S. stercoralis* with molecular taxonomic information, there is a strong sampling bias towards southeast and east Asia and Australia.

**Methodology/Principle findings:** We collected human and dog derived *Strongyloides* spp. and hookworms from two locations in Bangladesh and subjected them to molecular taxonomic and genomic analysis based on nuclear and mitochondrial sequences. All hookworms found were *Necator americanus*. Contrary to earlier studies in Asia, we noticed a rather high incidence of *S. fuelleborni* in human samples. Also in this study, we found the two types of *S. stercoralis* and no indication for genetic isolation from the southeast Asian populations. However, we found one *S. stercoralis* worm in a human sample that genomically was of the “dog only” type and we found two worms in a dog sample that had the nuclear genomes of the “dog only” type but the mitochondrial genome of the “human and dog” type.

**Conclusions/Significance:** *S. fuelleborni* may play a more prominent role as a human parasite in certain places in Asia than previously thought. The introgression of a mitochondria haplotype into the “dog only” population suggests that rare interbreeding between the two *S. stercoralis* types does occur and that exchange of genetic properties, for example a drug resistance, between the two types is conceivable.

**Author Summary:** More than 600 million people are infected with the nematode intestinal parasite *Strongyloides stercoralis*. Dogs can also carry *S. stercoralis*. In southeast Asia different genetic types that either infect only dogs or humans and dogs were described. *Strongyloides fuelleborni*, (normally found in old-world monkeys) can also infect humans, mainly in Africa. We collected *Strongyloides* spp. and hook worms, from humans and a dog in Bangladesh and analyzed their nuclear and mitochondrial genomes. All hookworms were *Necator americanus*, one of the two major human hookworm species. Contrary to the general believe that human infections with *S. fuelleborni* are extremely rare in Asia, we found multiple such cases, suggesting that *S. fuelleborni* plays a more important role as a human parasite than previously thought also in Asia.

We found the two expected genetic types of *S. stercoralis*. For the first time we found a genomically “dog only” type worm in a person and we found two worms with nuclear genomes of the “dog only” type but mitochondrial genomes of the “human and dog” type. This suggest that rare interbreeding between the two types occurs, such that exchange of genetic properties, such as a drug resistance, between the two types is conceivable.

## Introduction

Strongyloidiasis is one of the soil-transmitted helminthiasis (STH), which are recognized as neglected tropical diseases (NTDs, https://www.who.int/health-topics/neglected-tropical-diseases#tab=tab_1). However, although clearly more prevalent in tropical and sub-tropical areas, the disease is not limited to these regions and should probably be rather regarded as a disease of socioeconomically disadvantaged people rather than strictly a tropical disease [1, 2]. Until recently, strongyloidiasis was often neglected, even in comparison with other STHs [3] but over the last few years there is increasing interest in this disease [1, 4, 5]. The estimate of the number of people currently infected with *Strongyloides stercoralis*, the species which causes the vast majority of human strongyloidiasis cases, has recently been corrected upwards to “about 600 million” [2]. Given the difficulties with diagnosis, the true number may be even higher [6–8]. Although *S. stercoralis* has also been reported to occur in non-human primates (NHPs), dogs and cats, rendering these animals putative sources for zoonotic human *S. stercoralis* infection [6, 9–11]. In NHPs and in cats other, more or less host specific species of *Strongyloides* were also described, i. e. *S. fuelleborni* and *S. cebus* in NHPs, and *S. planiceps* and *S. felis* in cats [12]. The species status of *Strongyloides* in dogs has been controversially discussed ever since Brumpt separated the *Strongyloides* in dogs as *S. canis* from the human infective species *S. stercoralis* [13], interestingly based on the very same data that had convinced Fülleborn that the *Strongyloides* he found in the dogs belonged to the same species as the ones in humans [14]. While it became clear that dogs can be experimentally infected with at least some isolates of *S. procyonis* (natural host racoon, [15]) and human infective *S. stercoralis* (reviewed in [10]) it remained enigmatic if the species causing most natural *Strongyloides* infections in dogs is different from the one in humans.

Over the last years, the Hyper Variable Regions (HVR) I and IV of the nuclear Small SUbunit ribosomal RNA locus (*SSU*) and the mitochondrial *cox-1* loci have emerged as the standard markers for molecular taxonomy within the genus *Strongyloides* spp. and within the species, *S. stercoralis* [16–19]. A nomenclature system for the different haplotypes has been proposed and extended [18, 20, 21]. A few studies, analyzing samples from East and southeast Asia [20, 22–24], and Iran [25] analyzed whole genome data from individual *S. stercoralis* worms in addition to the *SSU* and *cox-1* markers. Jaleta et al. [20] and Nagayasu et al. [26] in southeast Asia and Beknazarova et al. in Australia [27] found that dogs carried at least two types of *Strongyloides*, only one of which appeared to be shared with humans in the same region. In this manuscript we will refer to them as “human and dog” and “dog only”, respectively. Barratt and Sapp [21] compiled all sequence information available from *S. stercoralis* from different hosts and used machine learning approaches to analyze these data in depth. Their findings suggest that *S. stercoralis* is in fact a complex of closely related species with different but overlapping host spectra. While from these data it appears most likely that zoonotic *S. stercoralis* infections can happen, currently, we do not know how important such infections are for the overall *S. stercoralis* epidemiology, compared with human to human transmission [10]. In addition to *S. stercoralis*, *Strongyloides fuelleborni*, which can be distinguished from *S. stercoralis* morphologically [28] and coprologicaly (from this species eggs are shed with the faeces and not larvae as in *S. stercoralis* [29]), has been found to be able to infect humans. Two subspecies of *S. fuelleborni* have been described, namely *S. fuelleborni fuelleborni* and *S. fuelleborni kellyi* [29]. While the former is the predominant species of *Strongyloides* in old world non-human primates, *S. fuelleborni kellyi* has been found only in humans in Papua New Guinea [29]. Based on molecular taxonomy [30] we think *S. f. kellyi* should probably be considered a separate species and do not further discuss it in this publication. For the rest of this publication “*S. fuelleborni”* always refers to *S. fuelleborni fuelleborni.* Barratt and Sapp [21] described genetic/genomic differences between *S. fuelleborni* in Africa and *S. fuelleborni* in Asia. The vast majority of human *S. fuelleborni* infections were found in Africa and [21] found genetic indication for a human specialized sub-population within the African clade, suggesting human to human transmission. In Asia, on the other hand, no such genetic hint was found and it appears that human *S. fuelleborni* infections are restricted to individuals with close contact to non-human primates, indicating that most, if not all human *S. fuelleborni* cases in Asia are zoonotic [21] and references therein.

So far, for *S. stercoralis* with molecular taxonomic information, there is a strong sampling bias towards southeast Asia, East Asia and Australia [21]. To further extend the geographic range, we collected *S. stercoralis* from two locations in Bangladesh. We knew that *S. stercoralis* is prevalent in Bangladesh ([2] lists an estimated overall prevalence of 17.3%), but we are not aware of any published systematic study on *S. stercoralis* in this country. Contrary to earlier studies in Asia, we noticed a rather high incidence of *S. fuelleborni* in humans. Molecular taxonomically these worms grouped clearly with the Asian clade defined by [21]. Molecular taxonomically, the *S. stercoralis*, we found mixed in with the southeast Asian population described earlier [20, 22, 23, 26] and appeared not to form a separate population. However, we found one *S. stercoralis* worm in a human sample that was of a type that had been considered dog-specific and we found two worms in a dog sample that had the nuclear genomes of the “dog only” type but the mitochondrial genome of the “human and dog” type, suggesting that occasional interbreeding between the types does occur.

## Methods

### Ethics statement

All participants were volunteers and gave informed consent. The sampling of human-derived material including the procedures to obtain informed consent, was in accordance with the Bangladeshi legal requirements and with the guidelines of the Sylhet Agricultural University. This study was approved by the Ethical Review Committee, Sylhet Agricultural University Research System (SAURES), Bangladesh (SAURES-UGC-2022-04). Interested putative participants were informed orally about the project and, if they chose to participate, were handed collection containers. All participants remained free to return the container or not.

### Study area, Sample collection and processing

Human faecal samples were collected from two regions from Bangladesh that had been previously identified as high prevalence areas for helminthiasis: Sylhet, and Dhaka in December 2022. 134 human samples from four different locations in Sylhet; Khadim tea garden, Daldali tea garden, Baluchar and Fotehpur and one dog sample from the premises of Sylhet Agricultural University were analysed. In Dhaka 95 human samples were collected from two locations: Hazaribug and Mohammadpur.

Stool collection jars with a spoon were distributed to the individuals who agreed to participate in the study after explaining how to properly collect the stool sample without soil contamination. The next day the sample jars were collected. Faecal samples were mixed well with approximately equal volumes of activated charcoal (Roth 5966.1) to facilitate air exchange. Water was added to make the samples well moisturized but not soakingly wet. This mixture was incubated in the room temperature (R.T.) for 24-48hrs with the lid partially open and re-moisturized on need-to basis. Samples were analysed using modified miniature Baermann apparatuses based on 50ml Falcon Tubes as described [31]. In brief, faeces mixture was placed in the centre of a 10×10cm cotton gauze and made it into a pouch secured using a toothpick which was submerged in a 50ml falcon tube filled with lukewarm water. After 3hrs of incubation at ambient temperature the sediment was taken out using a Pasteur pipette and observed under a stereo dissecting microscope for the presence of worms. These worms in part individually and in part bulk preserved in 80% ethanol at the Sylhet Agricultural University as described [32] and brought back to the Max Planck Institute for Biology, Tübingen for molecular/genomic analysis.

### Single worm lysis and *cox-1*, *SSU* HVR-I and *SSU* HVR-IV genotyping

Single worm lysis for adults and larvae was performed as described [32]. For infective larvae the lysis was extended to 6 hrs at 65°C. The lysate was either freshly used for PCR or stored at -20°C. The *cox-1*, *SSU* HVR-I and *SSU* HVR-IV were PCR amplified using the primers described in [24]. For the *SSU* HVR-I and the *SSU* HVR-IV for all worms the same primer pairs RH5401/RH5402 and 18SP4F/18SPCR, respectively, were used. For *cox-1* the primers designed for *S. stercoralis* (but also working for some other species) ( ZS6985/ZS6986) were used, unless the worm was already known to be a hookworm, in which case the hookworm optimized primer ZS6989 was used as the reverse primer instead of ZS6986. For the PCR, 10µl of QIAGEN *Taq* PCR master mix (x2) (201443), 0.4µl of 10µM forward and reverse primers, 7.2µl of PCR water and as template 2µl of single worm lysate were added. For *S. fuelleborni* the same primers as for *S. stercoralis* were used.

For sequencing 1µl of PCR product was mixed with 1µl of the relevant sequencing primer (10µM) [24] and 8µl of water and submitted to Genewiz, Leipzig, Germany. *S. stercoralis* P203 iL3 HVR-I PCR product and *S. fuelleborni cox-1* PCR products were gel purified (1% agarose, 1X TAE) using the QIAquick gel extraction kit (Qiagen 28706) prior to sequencing. If the sequencing result was not clean, the PCR products were sequenced again using the alternative sequencing primers listed by [24] or the amplification primers. Sequence quality and the presence of hybrid sequences was manually assessed by looking at the chromatograms using the SnapGene® software (from Dotmatrics; available at snapgene.com). The sequences were first compared with published sequences in the National Centre for Biotechnology Information (NCBI) database using the BLAST function (http://blast.ncbi.nlm.nih.gov/Blast.cgi) and then phylogenetically analysed using MEGA11 [33] with the Neighbour-joining method and default settings. The robustness of the trees was assessed with 1000 boot strap repetitions. Position numbers refer to GeneBank entries AF279916 for the *S. stercoralis* and *S. fuelleborni SSU*, LC050212 for the *S. stercoralis* and *S. fuelleborni cox-1* and AJ417719 for hookworm *cox-1* sequences.

We also retrieved *cox-1* sequences from the read data of the whole genome sequenced worms in [20] (see Fig. 4 of this reference) and submitted them to GenBank along with the *cox-1* sequences from this study (accession numbers OR804688-712, OR805174, 81, OR810937-54, OR809277-99).

### Whole genome sequencing

Whole genome sequencing was done as described [25] with slight differences as follows. In the DNA clean-up step, 11-14µl of lysis were used instead of 10µl. In the pooling and concentration adjustment step the bead clean-up was skipped. The concentrations were calculated for the samples and 2nM of each sample were pooled which resulted in a 1.85nM final concentration in the pool which was then submitted to the MPI for Biology in-house sequencing facility for Illumina NexSeq 2000 sequencing.

### Analysis of the whole genome sequences

#### Whole genome tree

The paired end sequencing resulted 5.7-21.5 x coverage for the *S. stercoralis* samples. WGS data for 12 *S. stercoralis* samples and three *S. fuelleborni* samples were uploaded to the European Nucleotide Archive under the study accession PRJEB70604.

The read alignment, duplicate removal, variant calling, defining heterozygous sites, creating the genotype using variant positions and constructing the NJ tree based on the variant positions were all done as described [25].

#### Whole mitochondrial (wmit) tree

WG sequencing reads from this study and previous studies were aligned to the *S. stercoralis* wmit reference genome (NC_028624.1 [identical to LC050212]) to generate wmit sequence assemblies. The four wmit sequences from [34] were used directly. Read alignment, binary alignment file generation and duplicate removal were done as mentioned above. BAM files were loaded to IGV_2.16.0 and the consensus sequences were obtained using the ‘copy consensus sequence function’ in IGV. NJ trees were generated using MEGA 11. The sequence file used as input for MEGA 11 can be found in S1 File.

To generate wmit assemblies from the *S. fuelleborni* in this study we first aligned the reads to two mitochondrial whole genome sequences from [35] (OL505577, arrangement A and OL602833, arrangement B) and visualized the BAM files using IGV. Since the tRNA(Met) gene that is present in arrangement B but absent in arrangement A was absent from all our *S. fuelleborni* sequences we decided to use OL505577 as reference. A wmit tree for *S. fuelleborni* was generated using sequences from this study and from [35] as described for *S. stercoralis* above.

#### Heterozygosity analysis

General heterozygosity analysis was done as described [25].

#### Coverage analysis

Coverage for both autosomal contigs and X-chromosomal contigs were analysed using samtools (0.1.18) depth command and the coverage was plotted against the number of positions using R studio.

## Results

We analysed 134 human samples (in most cases rather small samples) collected from Sylhet and found worms in 25 of them. In seven samples we found only *Strongyloides*, in five samples *Strongyloides* and hookworms, in 12 samples only hookworms and in one sample we found several worms that, based on their 18S sequence belonged to *Tokorhabditis* spp., which is a genus of free-living nematodes [36]. We think this last case represents a contamination from the ground and this sample is not further discussed. The one dog sample we obtained was positive for *Strongyloides.* From Dhaka, we found *Strongyloides* in only two out of 95 human samples, while, based on 18S sequence, we detected *Caenorhabditis nigoni*, which are free-living nematodes [37] in six of them. These worms likely represent ground contamination and are not discussed further. For 71 hookworms and 99 *Strongyloides* (67 from humans, 32 from the dog) the sequence of at least one out of the *SSU* HVR-I, *SSU* HVR-IV or *cox-1* was successfully determined.

### The hookworms found were *Necator americanus*

Initially 47 worms were confirmed to be hookworms, based on *SSU* sequences. Since the different hookworm species cannot be distinguished based on their *SSU* HVR-I or *SSU* HVR-IV sequences, we determined the *cox-1* sequence using the same primers used in [24], which was successful for 42 worms. Among these worms, we identified 18 different *cox-1* haplotypes (accession numbers OR810937-54), of which six were identical with existing database entries, while 12 were new (S1 Table). In several occasions worms with different *cox-1* sequences were found within the same host (S1 Table). All 18 sequences clustered with perfect bootstrap support with the group A [38] which is considered *Necator americanus* (Fig. 1). Within the species *N. americanus*, our samples did not cluster together but intermixed with sequences derived from Africa and Asia, arguing against the presence of a Bangladesh specific sub-population. In addition, 24 larvae for which we amplified the *cox-1* sequence using the primers optimized for *S. stercoralis* turned out to be hookworms. Since these sequences were shorter than the ones generated using the hookworm specific primers, they are not included in Fig. 1. However, all 24 sequences clearly grouped with *Necator americanus* sequences. Hence, overall, we identified 71 worms as hookworms, of which we confirmed 66 to be *Necator* and not *Ancylostoma*.

**Fig. 1:**
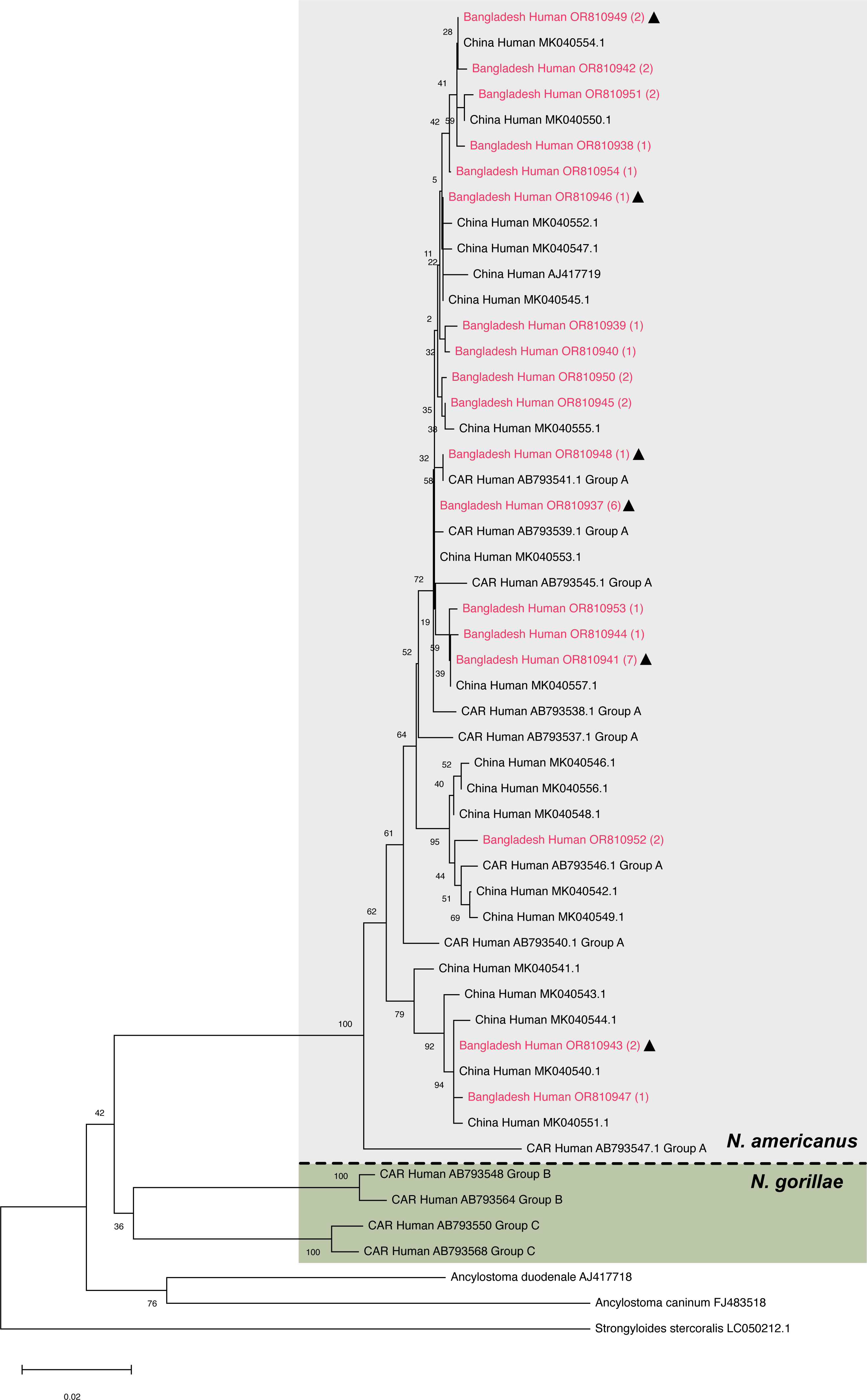
Neighbour joining tree based on partial hookworm *cox-1* sequences (670bp) . Sequences found in this study are in red, the number of worms this sequence was found in is in (). Triangles indicate haplotypes that had been previously known. For every sample the country of origin (CAR=Central African Republic), the host and the GenBank accession number are given.

### High incidence of *S. fuelleborni*

Strikingly, four out of the 12 individuals found to be infected with *Strongyloides* spp. in Sylhet carried *S. fuelleborni* and not *S. stercoralis* (two of them were co-infected with hookworms).

At the *SSU*, all 16 *S. fuelleborni* genotyped were HVR-I haplotype XIV and HVR-IV haplotype S (cf. [21]). Out of the 16 worms, *cox-1* sequences were obtained for 15. We identified eight different *cox-1* haplotypes (accession numbers OR805174-81), none of which had been reported before (S1 Table). All three persons for whom we obtained the *cox-1* sequence from more than one worm, carried worms with different haplotypes (S1 Table). In a phylogenetic analysis based on the *cox-1* sequences (Fig. 2), all eight sequences clearly grouped with *S. fuelleborni* from southeast Asia (mainly Thailand, Myanmar and Laos) described as cluster 3 by [21], further supporting the notion that in *S. fuelleborni* (different from *S. stercoralis*) geographic sub-populations exist and an Asian and an African clade exist [21].

**Fig. 2:**
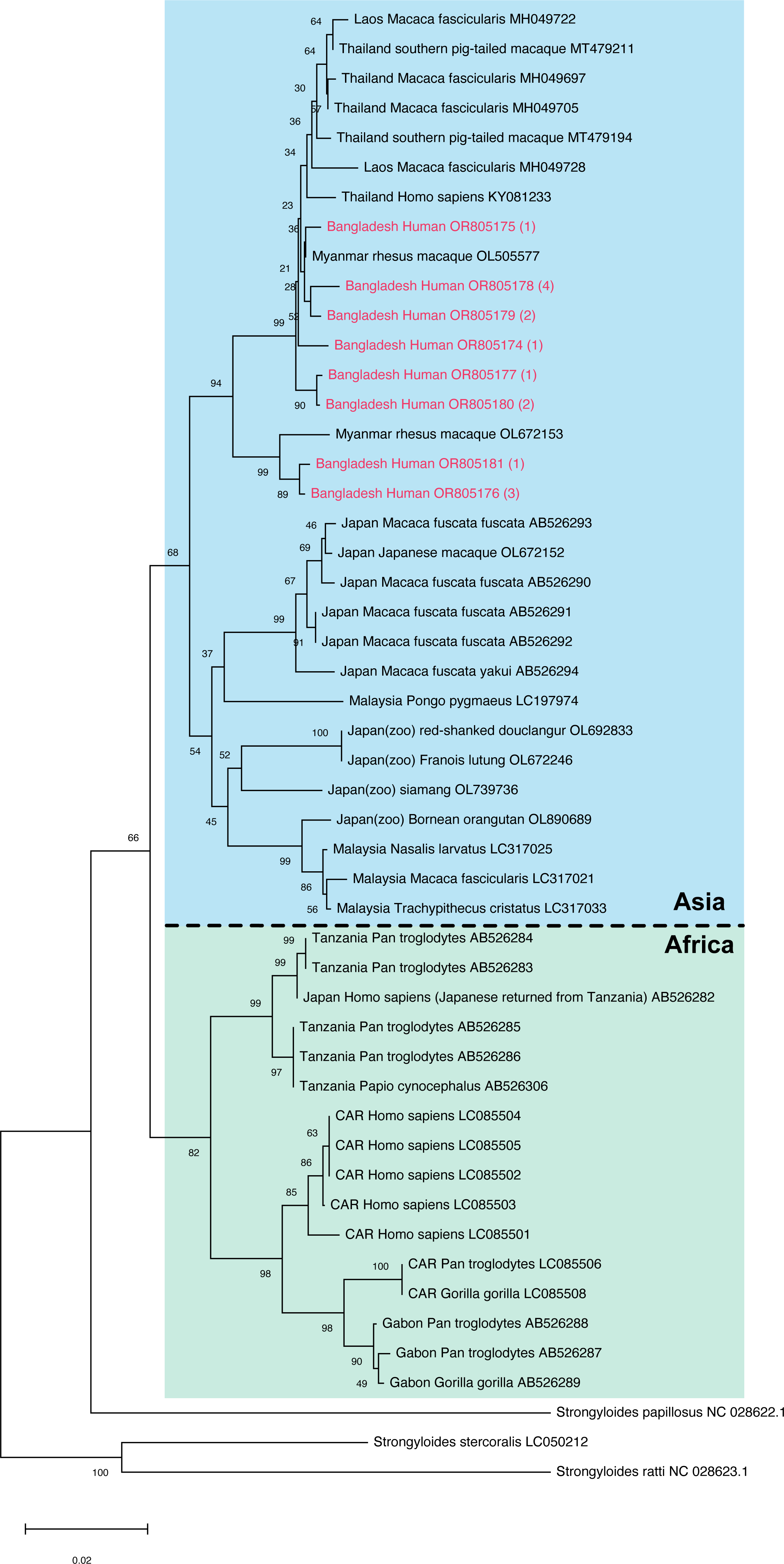
Neighbour joining tree based on partial *S. fuelleborni cox-1* sequences (552bp) . Sequences found in this study are in red, the number of worms this sequence was found in is in (). For every sample the country of origin (CAR=Central African Republic), the host and the GenBank accession number are given.

From three *S. fuelleborni* derived from three different persons we performed whole genome Illumina short read sequencing (Table 1) and deposited the read data in the European Nucleotide Archive (accession number PRJEB70604). Since there is no reference nuclear genome for this species available, we make these data publicly available here without further analysis. From these data we extracted the full mitochondrial genomes and compared them with the sequences reported by [35]. The sequences analysed are listed in S1 File. All three worms clustered with the samples containing mitochondrial genome arrangement A and did not contain the tRNA(Met) gene that is absent from arrangement A but present in arrangement B. Arrangement A had been observed in *S. fuelleborni* derived from macaques in Myanmar and Japan and was hypothesized to represent the ancestral state [35] (Fig. 3).

**Table 1:** Whole genome sequenced samples.

| Sample number | Worm ID | Species | *cox-1* haplotype (type) | *SSU* HVR-I haplotype | *SSU* HVR-IV haplotype | Internal ID in sequencing protocol |
| --- | --- | --- | --- | --- | --- | --- |
| 1 | Human_C19 - f2 | *S. stercoralis* | OR804711 | II | A | 1 |
| 2 | Human_C19 - f4 | *S. stercoralis* | OR804711 | II | A | 3 |
| 3 | Human_F40 - f1 | *S. stercoralis* | OR804710 | II | A | 14 |
| 4 | Human_F10 - f1 | *S. stercoralis* | OR804710 | II | A | 17 |
| 5 | Human_F10 - f3 | *S. stercoralis* | OR804709 | II | A | 19 |
| 6 | Human_J102 - f1 | *S. stercoralis* | OR804706 | I | U | 20 |
| 7 | Dog - f1 | *S. stercoralis* | OR804688 | I | U | 21 |
| 8 | Dog - f2 | *S. stercoralis* | OR804689 | I | U | 22 |
| 9 | Dog - f4 | *S. stercoralis* | OR804691 | I | U | 24 |
| 10 | Dog - f5 | *S. stercoralis* | OR804692 | I | U | 25 |
| 11 | Dog - f6 | *S. stercoralis* | OR804693 | I | U | 26 |
| 12 | Dog - m2 | *S. stercoralis* | OR804701 | I | U | 27 |
| 13 | Human_A22 - f1 | *S. fuelleborni* | OR805174 | XIV | S | 29 |
| 14 | Human_G22 - f1 | *S. fuelleborni* | OR805178 | XIV | S | 33 |
| 15 | Human_V132 - f1 | *S. fuelleborni* | OR805181 | XIV | S | 36 |

**Fig. 3:**
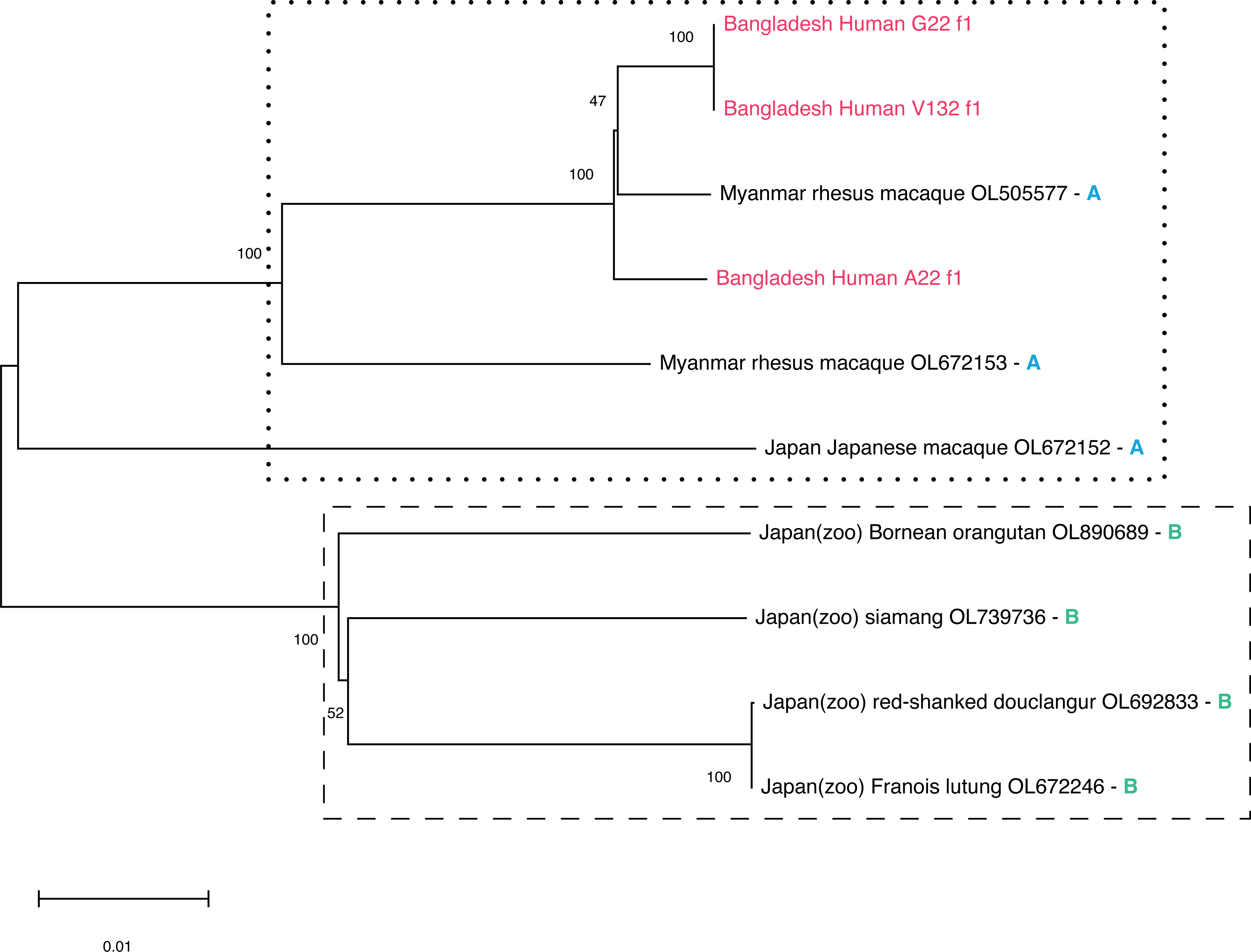
Neighbour joining tree based on full mitochondrial nucleotide sequences. All non-Bangladesh sequences are from Ko et al. (2023). The Letters (A or B) refer to the genome arrangement described in **Ko** et al. (2023) (compared with B, arrangement A lacks a tRNA(Met) gene present adjacent to the *S1* gene). For every sample the country of origin, the host and the GenBank accession number or the worm identifier (for sequences from this study) are given. The sequences from this study were extracted from the data available from the European Nucleotide Archive under the accession number PRJEB70604. A FASTA file with all sequences used is provided as S1 File.

### S. stercoralis SSU haplotypes

For *SSU* HVR haplotype nomenclature see [21]. At the *SSU* HVR-IV the *S. stercoralis* in our samples had either haplotype A (28 individuals), which was described to be indicative for the “human and dog” type [20, 21] or a new haplotype we call U (31 individuals, Fig. 4a). All but one (see below) of the carriers of haplotype U were isolated from the dog. At *SSU* HVR-I, 28 worms had haplotype I, 26 worms had haplotype II and one worm each had haplotype III and V. In all cases where we have the sequences for both HVRs, HVR-IV haplotype A co-occurred with HVR-I haplotype II (24 cases) or III (one case) while HVR-IV haplotype U co-occurred with HVR-I haplotype I (28 cases) or V (one case). Notice that in an earlier study in Cambodia [20], HVR-I haplotype I did also co-occur with HVR-IV haplotype A.

**Fig. 4:**
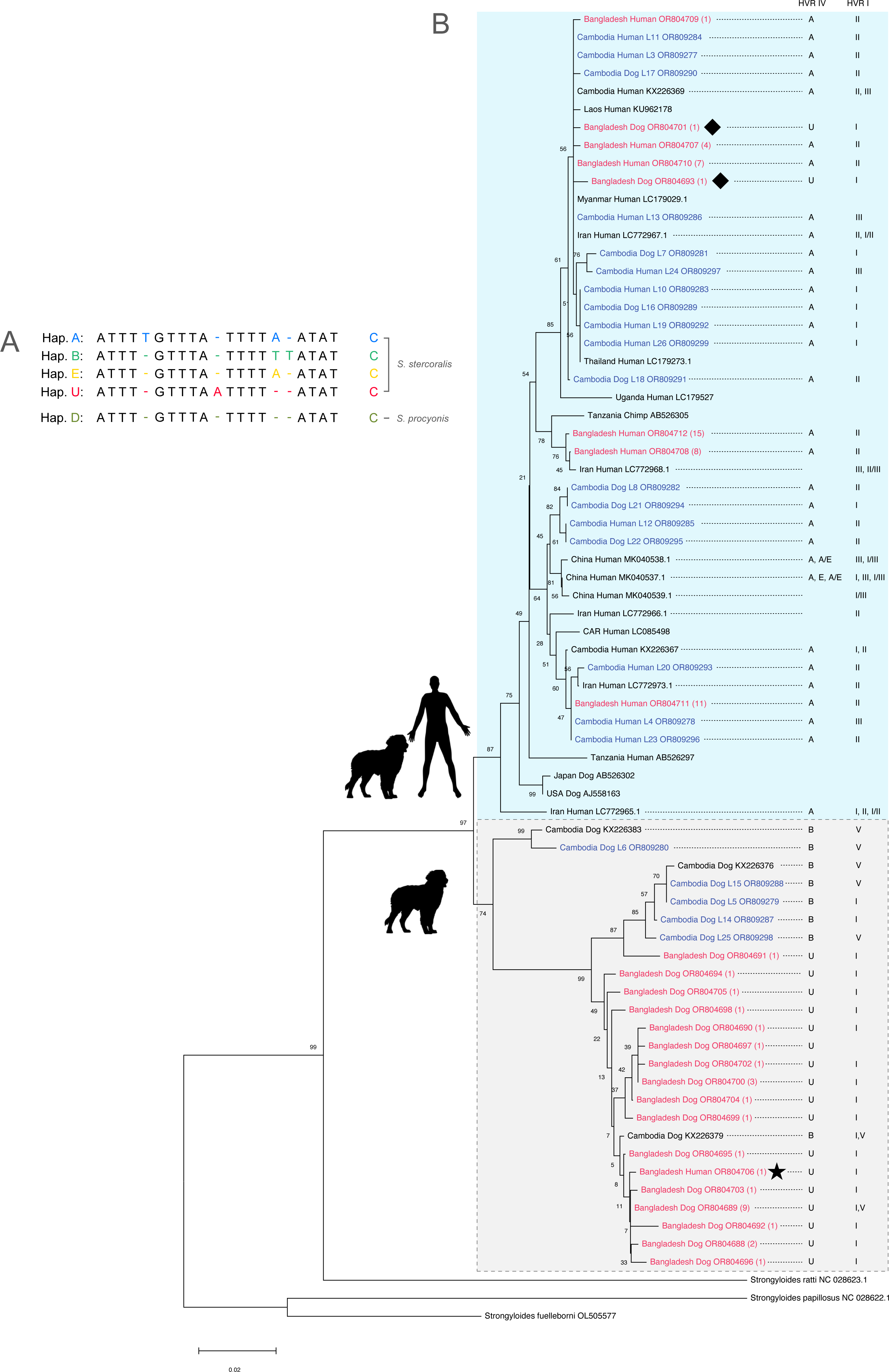
*cox-1, SSU* HVR-I and *SSU* HVR-IV haplotypes of our samples compared with selected published sequences. A: sequence of the new *SSU* HVR-IV haplotype U compared with the previously reported haplotypes (nomenclature according to [21]) mentioned in B. Notice that haplotype E was called haplotype C in Zhou et al. (2019). B: neighbour joining tree based on partial *S. stercoralis cox-1* sequences (552bp). Sequences found in this study are in red, the number of worms this sequence was found in is in (). Samples in blue are worms from [20] for which full genome short read sequences are available. For every sample the country of origin (CAR=Central African Republic, USA=United States of America), the host and the GenBank accession number is given. For worms from Cambodia also the worm individual is given after the host to facilitate cross reference with [20]. For each *cox-1* haplotype the *SSU* HVR-I and *SSU* HVR-IV haplotypes found in individuals with this *cox-1* haplotype are indicated (if known). Samples from Cambodia are from [20], samples from Laos are from [39], samples from Myanmar, Thailand and Uganda are from [26], samples from Iran are from [25], sample from the USA is from [40], samples from Tanzania, and Japan are from [17], samples from CAR are from [41] and samples from China are from [24]. Haplotypes separated by ‘/’ indicates a heterozygous worm. For a *cox-1* tree with more sequences see S1 Fig. Diamonds label the two worms that showed “human and dog” type mitochondrial but “dog only” type nuclear sequences. The asterisk labels the worm in the “dog only” cluster isolated from a human host.

### S. stercoralis cox-1 haplotypes

In a total of 76 worms, we detected 25 different *cox-1* haplotypes (accession numbers OR804688-712) of which three had been previously reported while 22 were new (S1 Table). Each haplotype was present in between one and 15 different worms with the previously known haplotypes being the first (15 worms), second (11 worms) and fifth (seven worms) most abundant ones (Fig. 4). Upon phylogenetic analysis (Fig. 4, S1 Fig) the previously described and five of the new haplotypes (representing 48 worms, two of which had been isolated from the dog) grouped with sequences in the “human and dog” clusters according to [20]. 17 haplotypes (representing 28 worms, of which one had been isolated from a human sample [see below]) grouped with one of the “dog only” clusters according to [20].

### A “dog only” type *S. stercoralis* in a human host

Strikingly, one of the worms that by *cox-1* sequence fell into the “dog only” cluster had been isolated from a human from Dhaka (worm: Human_J102 - f1, asterisk in Fig. 4B). This was the sole worm found in this particular sample and it carried the new *SSU* HVR-IV haplotype U, and HVR-I haplotype I, like all but one (the one with HVR-I haplotype V) of the other worms from our study in this cluster, which all came from the dog.

### First finding of a “human and dog” type mitochondrial genome in combination with a “dog only” type nuclear genome

In all previous studies that we are aware of, where nuclear and mitochondrial sequences from the same worms were determined [20, 23, 26, 42, 43] both sequence kinds always fell in the same group as defined by [20, 26] (“human and dog” or “dog only”). Here, for the first time, we found worms where the phylogenetic positioning based on the *cox-1* and on the *SSU* sequences was not in agreement. In the dog we found two worms, that based on their *cox-1* sequences fell within the “human and dog” type but had *SSU* haplotypes normally associated with the “dog only” type (diamonds in Fig. 4B). This suggests that rare interbreeding between the two types does occur.

### The *S. stercoralis* population in humans is genetically close to the one in southeast Asia

Since the conclusions above are based on a rather small number of informative positions, we performed Illumina whole genome sequencing of the three notable worms mentioned above along with nine other individuals isolated from humans and the dog (Table 1). Then we compared their mitochondrial and nuclear genomes. The read data are available from the European Nucleotide Archive (accession number PRJEB70604). The extracted whole mitochondrial genomes are listed in S2 File.

We reconstructed Neighbour Joining cladograms based on the full mitochondrial (Fig. 5, S2 Fig) and nuclear (Fig. 6, S3 Fig, S4 Fig) genomes. In all cases the classification as “human and dog” or “dog only” agreed with the one based on only *cox-1* for the mitochondrial genome (cf. Figs. 5 and 4B) or only the *SSU* HVR-IV for the nuclear genome (cf. Figs. 6B and 4B). Notice that the nuclear genome tree should not be interpreted as a phylogenetic tree because it is a within-species tree with genomes possibly undergoing mixing due to meiotic recombination. Neighbour Joining clustering of the “human and dog” type whole nuclear genome sequences showed that the worms from Bangladesh group with the sequences previously described for southeast Asia, Japan and Iran and away from a possibly asexual population described in southern China and the laboratory reference isolate, which originated from the USA (Fig. 6A, for references see figure legend).

**Fig. 5:**
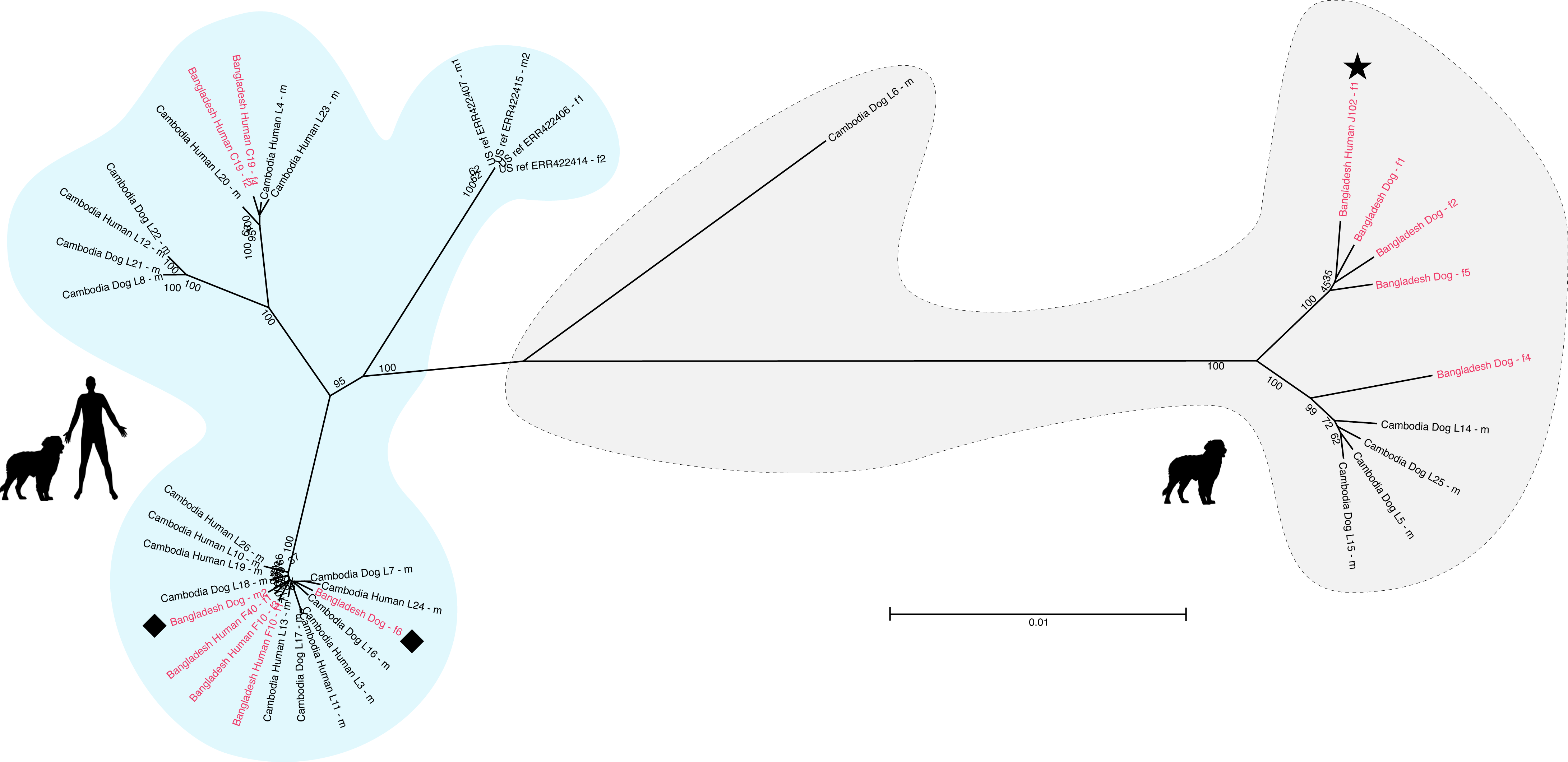
Neighbour joining tree based on full mitochondrial genomes determined in this study (in red) plus the worms mentioned in Fig. 4 of [20] and four worms of the reference isolate [44] for comparison. For a tree with more sequences see S3 Fig. A FASTA file with all sequences used in this figure and in S3 Fig is provided as S2 File. Diamonds label the two worms that showed “human and dog” type mitochondrial but “dog only” type nuclear sequences. The asterisk labels the worm in the “dog only” cluster isolated from a human host.

**Fig. 6:**
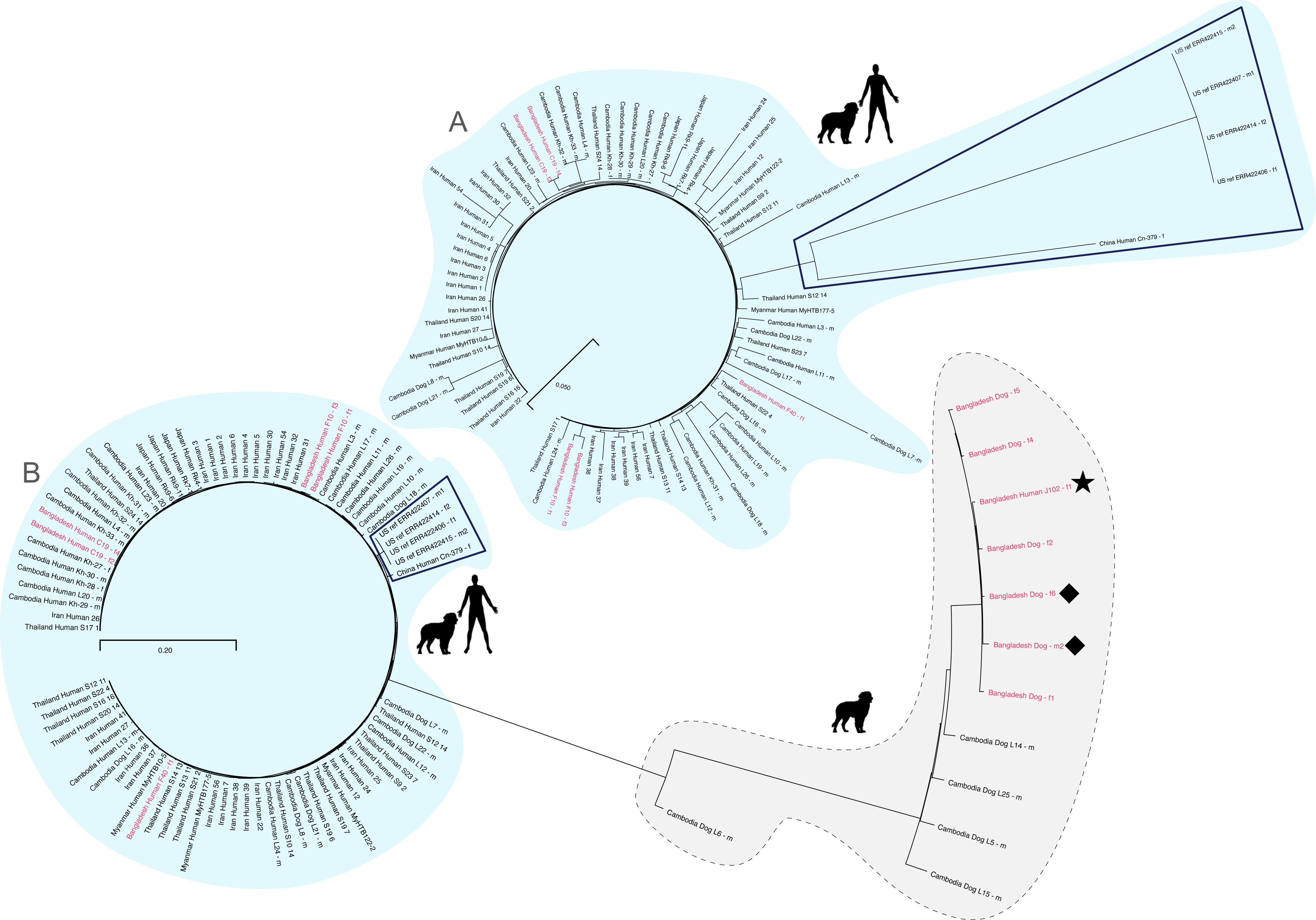
Neighbour joining tree (A without and B with “dog only” type worms) based on full nuclear genomes determined in this study (in red) plus selected worms from earlier studies for comparison. Diamonds label the two worms that showed “human and dog” type mitochondrial but “dog only” type nuclear sequences. The asterisk labels the worm in the “dog only” cluster isolated from a human host. To get an impression on how different the “dog only” type is from the “human and dog” type, compare in A and B the branch lengths of the samples from the USA and China (boxed), which are the “human and dog” type worms with the greatest genomic difference from the southeast Asian human derived *S. stercoralis.* For a different representation, see S3 Fig and S4 Fig. Sequences from Cambodia are from [20], Thailand are from [22], Iran are from [25], Myanmar and Japan are from [23], the USA are from [44] and China is from [24].

### Whole genome sequence confirms the unexpected worms

When the sequences of the “dog only” cluster are included in the analysis, there remains little resolution in the “human and dog” branch of the nuclear genome tree (Fig. 6B, S2 Fig). This is because, compared with the “dog only” sequences they are very similar to each other and the inclusion of more samples reduced the number of informative sites included in the analysis (only positions covered in all worms in the analysis were considered, see Materials and Methods). However, this analysis confirms the unexpected results based on the *SSU* and the *cox-1* sequences. Worm Human_J102 - f1 (isolated from a human host, marked with an asterisk in Figs. 5 and 6) grouped with respect to the nuclear and with respect to the mitochondrial genome with the “dog only” cluster, while worms Dog_f6 and Dog_m2 (marked with diamonds in Figs. 5 and 6) both show a mitochondrial genome that groups with the “human and dog” cluster but a nuclear genome that belongs to the “dog only” cluster. We conclude from this result that occasional interbreeding of the two types does occur and thereby a “human and dog” type mitochondrial genome introgressed into the “dog only” population. Presumably, a “human and dog” type female and a “dog only” type male interbred and the descendants later bred with “dog only” type partners, thereby rendering the recombining nuclear genome “dog only” type while maintaining the uni-parentally (maternally) inherited mitochondrial genome.

### Worms belonging to the “human and dog” type show low heterozygosity

In order to compare our samples from Bangladesh with earlier studies [22–25] we performed a heterozygosity analysis (Fig. 7A). The worms of the “human and dog” type from Bangladesh showed very similar heterozygosity like the ones described from southeast Asia [22, 23] and a portion of the worms from a recent study in Iran [25]. The heterozygosity was clearly lower than in the presumably essentially asexual populations in Japan [23] and in southern China [24] and in a portion of the worms from Iran [25].

**Fig. 7:**
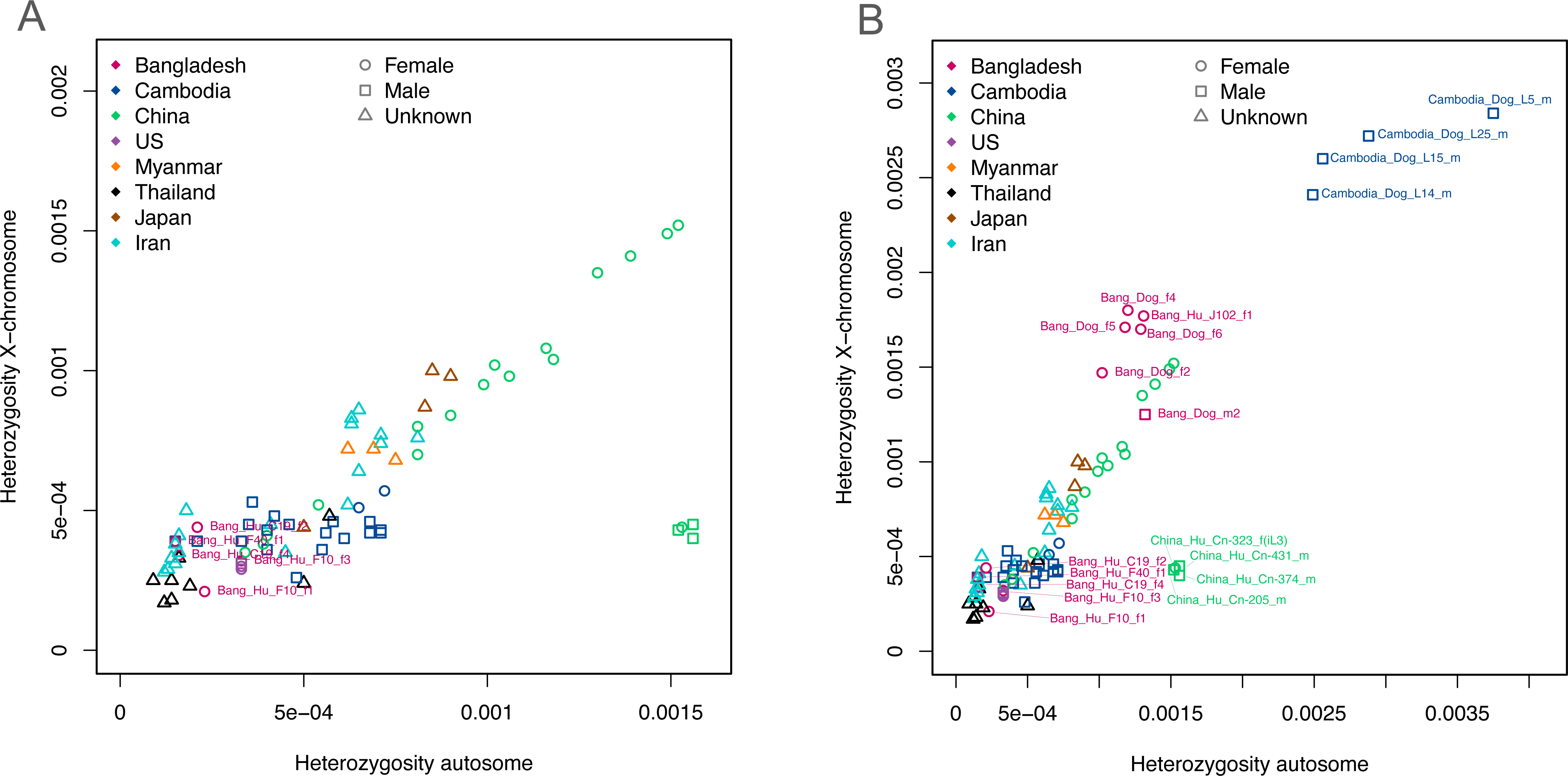
Measured heterozygosity of the whole genome sequenced worms from this and previous studies. The X axis shows the heterozygosity on the autosomes and the Y axis shows the heterozygosity on the X chromosome. A: only “human and dog” type worms. B: the same worms as in A plus the “dog only” type worms from this study and from [20]. Notice the high heterozygosity on the X chromosome in males of the “dog only” type. The samples are from previous studies are from the following references: Thailand [22], Iran [25], USA [44], Japan and Myanmar [23], Cambodia [20] and China [24].

### Worms belonging to the “dog only” type show high apparent heterozygosity that is likely caused in part by structural variations, rather than true heterozygosity

When we attempted to include the “dog only” type worms (including the one isolated from a human host) in the heterozygosity analysis we noticed that they showed very high apparent heterozygosity (Fig. 7B). Strikingly, this was also the case for heterozygosity on the X chromosome in males. To determine if this was an anomaly of our samples from Bangladesh we subjected the sequences of the five whole genome sequenced “dog only” type worms from [20] to the same analysis. Except for the worm L6, these sequences showed even higher apparent heterozygosity, including on the X chromosome (all “dog only” type whole genome sequenced individuals by [20] were males). Notice that with respect to the mitochondrial and the nuclear genomes, L6 belongs to a separate sub-cluster of the “dog-only” cluster than the other four worms and the worms isolated in this study. It is not clear, if worms in this subcluster are more closely related to the other “dog only” worms or to the “human and dog” worms (compare the positions of L6 in Figs. 4-6 and Suppl. Figs. 1,2). We think this high apparent heterozygosity is in part a consequence of using a divergent reference genome sequence (the reference sequence for the human infective *S. stercoralis* belongs to ‘human and dog’ cluster) for calling the heterozygous positions. The argument for this with Figs. is provided in S3 File and briefly summarized here. First, we asked if some of the *S. stercoralis* in Asia, in particular the “dog only” type might not employ XX/XO sex determination as it is the case in the USA derived *S. stercoralis* reference isolate [44]. We therefore performed read coverage analysis for males and females. Overall, in both types the X chromosome showed lower read coverage in males compared with autosomes and with females, suggesting that males of the “dog only” type do only have one X chromosome. We then analysed the heterozygosity over the length of the chromosomes. Males did indeed show very low heterozygosity over large portions of the X chromosome but there were apparent heterozygosity hot spots. These were visible in both males and in females and also on autosomes. We think these apparent heterozygosity hotspots reflect duplications and X to autosome translocations in the genome of the “dog only” type, compared with the *S. stercoralis* reference genome [44].

## Discussion

Among 12 *Strongyloides* positive persons from the Sylhet region we found four who carried *S. fuelleborni* rather than *S. stercoralis*. This was unexpected, since in Asia infections of humans with *S. fuelleborni* are considered very rare and restricted to people with very close interactions with monkeys [45–47].

In the Sylhet region there is a large population of free roaming monkeys. Albeit we have no reason to distrust out study participants, we need to point out that we cannot formally exclude that monkey faeces instead of human stool was returned. In future studies intending to confirm this ‘higher than expected’ infection of humans with *S. fuelleborni,* an independent confirmation of the host (e.g. through the detection of host specific sequences in the stool), would be desirable in order to dispel all doubts. Based on the *cox-1* sequences, the *S. fuelleborni* in Bangladesh were most closely related with worms from Thailand, Myanmar and Laos (cluster 3 in [21]) which makes sense due to geographic proximity.

As expected, we also found hookworms. Based on their *cox-1* sequences they were of the species *Necator americanus* (group A in [48]). In agreement with earlier studies [24, 48], we found no indication for population separation between Asia and Africa.

Overall, at the level of the nuclear and the mitochondrial genomes, the *S. stercoralis* we found in Bangladesh in humans mixed in with the worms described earlier from southeast Asia. Hence, we have no reason to assume that *S. stercoralis* in humans from Bangladesh and from southeast Asia are genetically distinguishable sub-populations. Together with the recent findings of [25] that *S. stercoralis* in Iran also share much of their genetic diversity with the ones in southeast Asia, these findings support the proposal by [26] and [34] that *S. stercoralis* has only rather recently established in humans after a host switch of a particular genotype from a canine host (possibly upon domestication of dogs) and then spread in the human population. It should, however, be noticed that based on reviewing published *cox-1* sequences, [19] did detect significant population structure and based on whole genome sequence, a possible asexual population of *S. stercoralis* in southern China and the laboratory reference isolate that originates from the USA, are genomically rather different from the southeast Asian *S. stercoralis* [24].

We found only one *Strongyloides* positive dog and all worms we analysed from this host individual had nuclear genomes that fell into the “dog only” cluster, based on the nuclear *SSU* and (if determined) whole genome sequences) (cf. [20, 21, 26]). *S. stercoralis* of the “dog only” type (based on molecular taxonomy) had so far been described only in southeast Asia [20, 26] and Australia [27] such that our findings extend the range, in which this type is known to occur, further West. The “dog only” type worms in this study showed a new *SSU* HVR-IV haplotype (now called haplotype U) that differs by one nucleotide from haplotype D (c.f. [21]). The fact that based on whole genome neighbour joining clustering all our dog derived worms grouped together with perfect bootstrap support should not be overinterpreted given that these worms were all derived from the same host individual and therefore might have been closely related. It is, however, noteworthy that the one worm that was isolated from a human but appeared genomically to belong to the “dog only” type also grouped with the dog derived worms although it had been isolated from Dhaka while the dog had been sampled in Sylhet. Again, given an N of one, this should not be over interpreted. In this study we made two unexpected observations. First, one of the human derived worms belonged genomically to the type that was so far considered to occur only in dogs. This one “dog only” type worm in a human does not invalidate the general conclusion about species specificity by [20, 21, 26]. Occasional zoonotic infections of humans with animal parasitic nematodes have been observed before, for example with filarial nematodes [49–53] or, even rather frequently, with animal parasitic hookworms [54, 55] (and references therein). It is therefore not really astonishing that with increasing sampling such a case emerged also for *Strongyloides*. We must point out that also in this case, we do not have an independent host confirmation and can therefore not exclude with absolute certainty that dog faeces was returned. Second and more importantly, we found two worms in the dog with nuclear genomes of the “dog only” type but mitochondrial genomes of the “human and dog” type. This was rather astonishing because, so far, in all cases where nuclear and mitochondrial sequences from the same worms had been determined [20, 23, 26, 42, 43] both sequence kinds always fell in the same cluster as defined by [20, 26] (“human and dog” or “dog only”). This finding suggests that at least occasional interbreeding of the two types does occur. A rare productive mating between a “human and dog” type female and a “dog only” type male followed by breeding with “dog only” type partners may have let to the introgression of the “human and dog” mitochondrial haplotype into the “dog only” population.

We found a very high apparent heterozygosity in worms of the “dog only” type. We think that this is in part an artifact caused by using the *S. stercoralis* reference genome, which is derived from a human infective isolate [44]. Compared with the reference, the “dog only” type, which, in our opinion, is likely to be a different species, might have a number of duplications with slightly deviating sequences. The positions that differ between the copies will be falsely considered heterozygous positions when the sequencing reads are aligned to the reference sequence without the duplication. Further, there might be translocations that are X chromosomal in the reference but autosomal in the “dog only” type. We think these findings illustrate that the two types are genomically rather different and that the *S. stercoralis* reference sequence is not suitable as a reference for certain genomic analyses of at least some of the “dog only” type *S. stercoralis*.

## Data Availability

All data are available in the manuscript, the supplementary materials or public databases under the accession numbers provided in the manuscript.

## Acknowledgements

We are grateful for infrastructural, logistic and practical field support to Hyeonmo Kim (International Parasite Research Bank), Tarek Siddiki, Proloy Chakraborty, Mohammad Rokibul Hasan Shanto (Sylhet Agricultural University), Hamida Khanum, Mandira Mukutmoni, Priyanka Barua, Ayan Goswami and Nusrat Jahan Moonmoon (University of Dhaka). We thank Keeseon Eom and the International Parasite Research Bank and the *Caenorhabditis elegans* and Nematode Bank at the Chungbuk National University for organisational support and the MPI for Biology Tübingen Genome centre for assistance with whole genome sequencing.

## Supporting information Captions

S1 File: FASTA file of the *S. fuelleborni* whole mitochondrial sequences used in Fig. 3.

S2 File: FASTA file of the *S. stercoralis* whole mitochondrial sequences used in S2 Fig (contains all sequences used in Fig. 5).

S3 File: Full argument for result section 2.10 Worms belonging to the “dog only” type show high apparent heterozygosity that is likely caused in part by structural variations, rather than true heterozygosity (text and three figures).

S1 Table: Excel table showing the different *cox-1* haplotypes for the *S. stercoralis, S. fuelleborni* and *N. americanus* found in this study in different tabs. The last tab shows the sequences extracted from the *S. stercoralis* whole genome sequencing read data from [20].

S1 Fig: NJ tree based on *cox-1* with more sequences compared with Fig. 4. Diamonds label the two worms that showed “human and dog” type mitochondrial but “dog only” type nuclear sequences. The asterisk labels the worm in the “dog only” cluster isolated from a human host. Sequences found in this study are in red, the number of worms this sequence was found in is in (). Samples in blue are worms from [20] for which full genome short read sequences are available. For every sample the country of origin (CAR=Central African Republic, USA=United States of America), the host and the GenBank accession number is given. For worms from Cambodia also the worm individual is given after the host to facilitate cross reference with [20]. Samples from Cambodia are from [20], samples from Laos are from [39], samples from Myanmar, Thailand and Uganda are from [26], samples from Iran are from [25], sample from the USA is from [40], samples from Tanzania, and Japan are from [17], samples from CAR are from [41] and samples from China are from [24].

S2 Fig: NJ tree based on whole mitochondrial genome sequences with more sequences compared with Fig. 5. Diamonds label the two worms that showed “human and dog” type mitochondrial but “dog only” type nuclear sequences. The asterisk labels the worm in the “dog only” cluster isolated from a human host. The sequences from this study are in red. The other sequences were extracted from the whole genome sequencing read data of the following references: Cambodia [20], Thailand [22], Iran [25], Myanmar and Japan [23], USA [44] and China [24]. A FASTA file with all sequences used is provided as S2 File.

S3 Fig (without the “dog only” cluster) and S4 Fig (including the “dog only” cluster): Different representation of the cladograms based on whole genome sequences compared with Fig. 6. Diamonds label the two worms that showed “human and dog” type mitochondrial but “dog only” type nuclear sequences. The asterisk labels the worm in the “dog only” cluster isolated from a human host. The sequences from this study are in red. The other sequences were extracted from the whole genome sequencing read data of the following references: Cambodia [20], Thailand [22], Iran [25], Myanmar and Japan [23], USA [44] and China [24].

## Notes

### Competing Interest Statement

The authors have declared no competing interest.

### Funding Statement

This study was funded by the Max Planck Society (Max Planck Institute for Biology Tuebingen, core budget).

### Author Declarations

This study was approved by the Ethical Review Committee, Sylhet Agricultural University Research System (SAURES), Bangladesh (SAURES-UGC-2022-04)

