## Supplementary figures and images for "Genomic analysis of Strongyloides stercoralis and Strongyloides fuelleborni in Bangladesh"

### S1 Fig

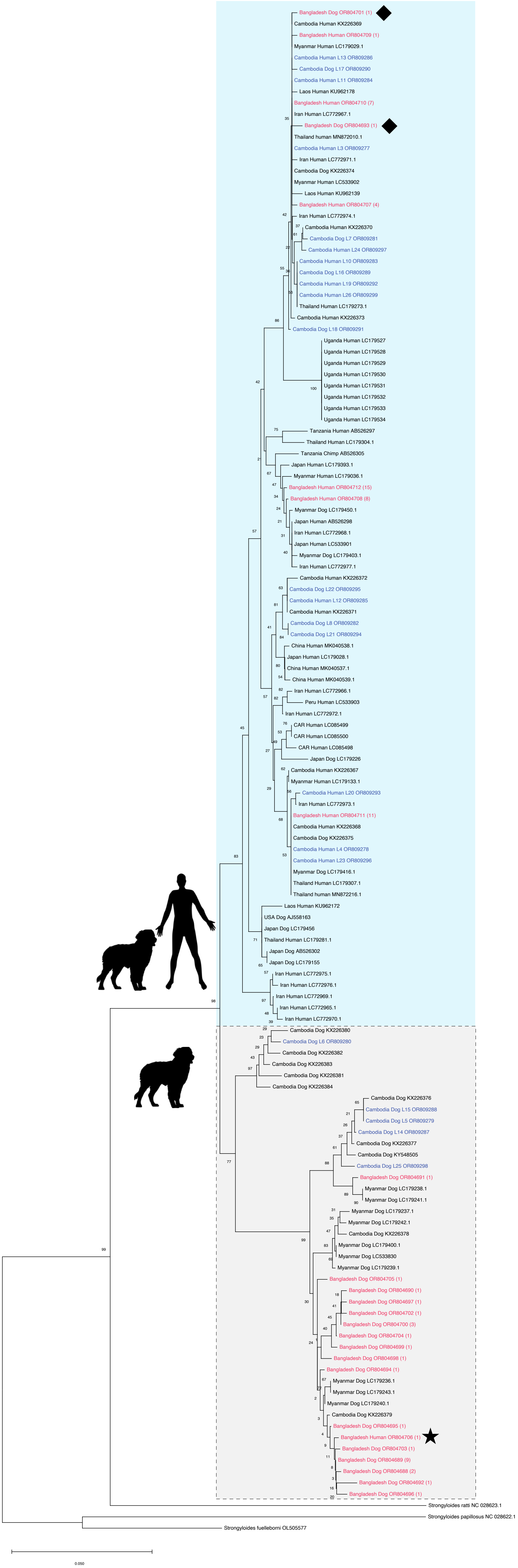

### S2 Fig.

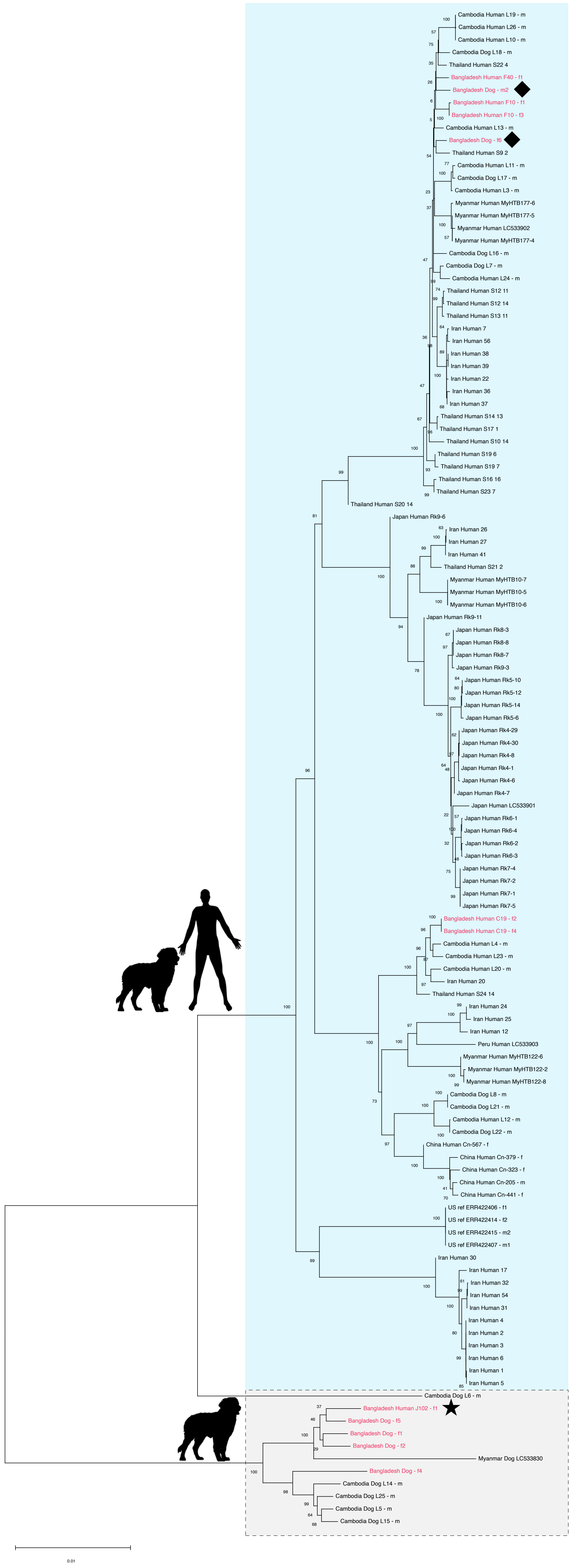

### S3 Fig.

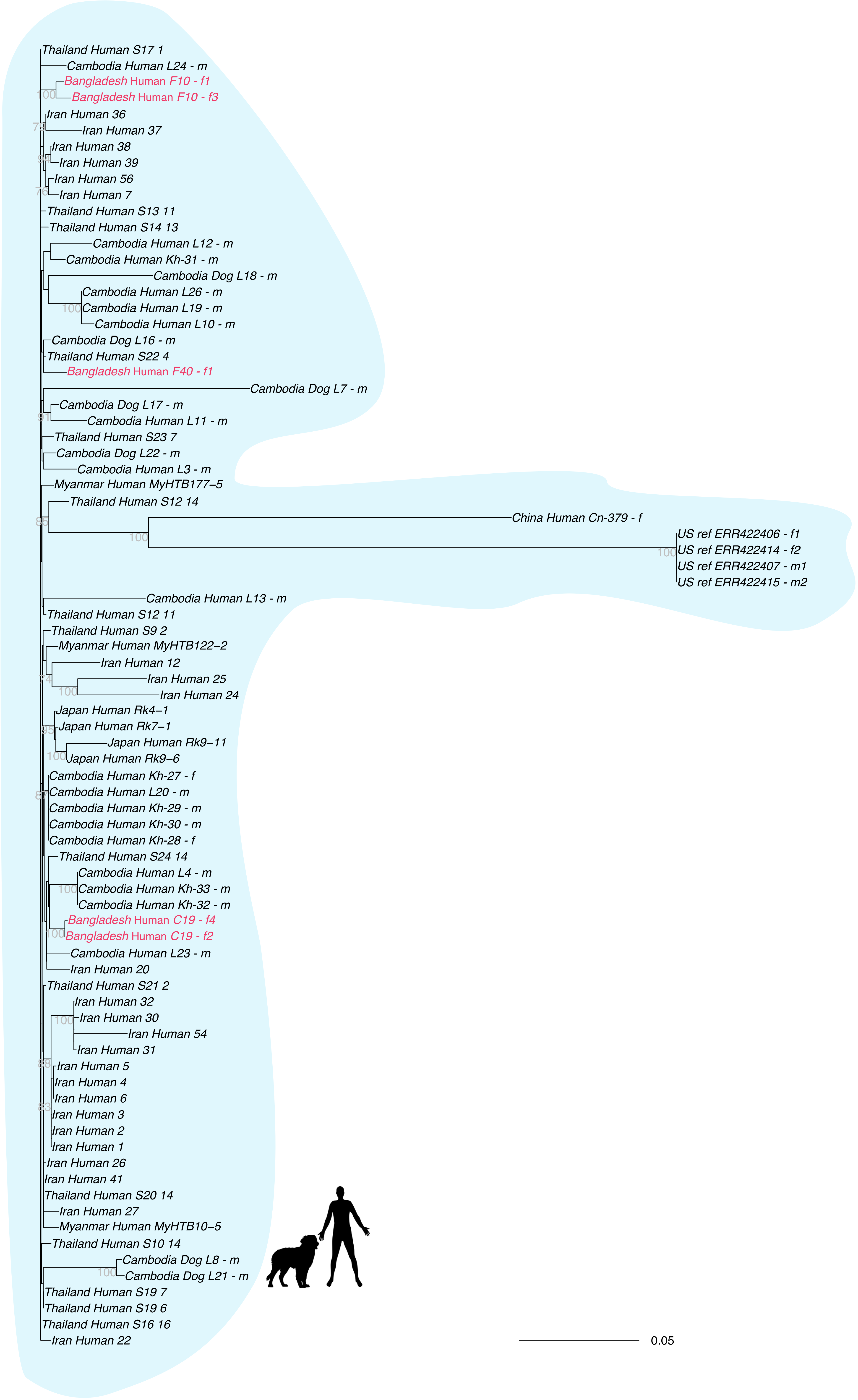

### S4 Fig.

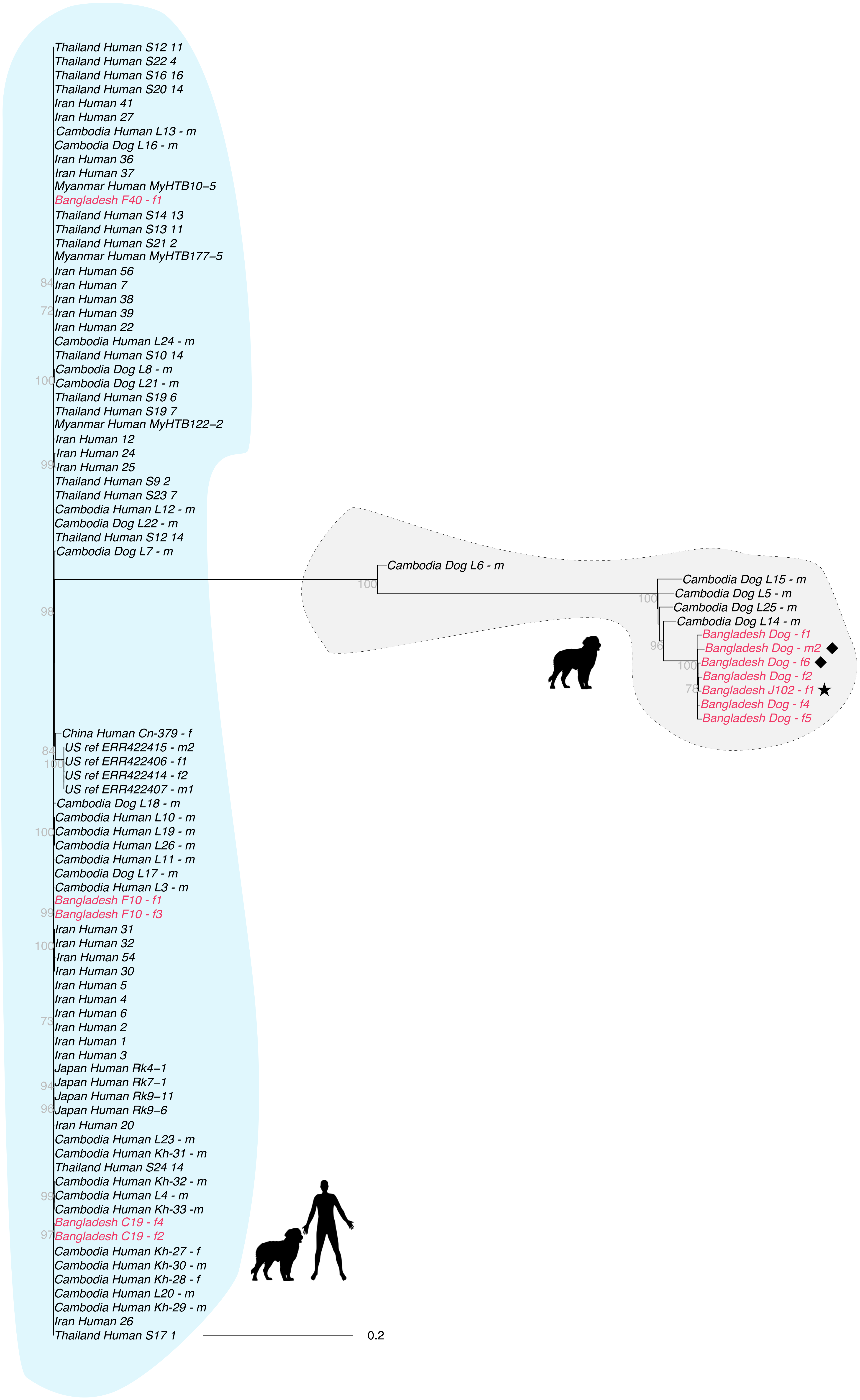
