## Supplementary material for "Genomic analysis of Strongyloides stercoralis and Strongyloides fuelleborni in Bangladesh": S3 File

**Supplementary File 3 to**

### Genomic analysis of *Strongyloides stercoralis* and *Strongyloides fuelleborni* in Bangladesh

Veroni de Ree[1], Tilak Nath[2], Dorothee Harbecke[1], Dongmin Lee[3], Christian Rödelsperger[1] and Adrian Streit[1]*

**Worms belonging to the "dog only" type show high apparent heterozygosity that is likely caused in part by structural variations, rather than true heterozygosity**

When we attempted to include the "dog only" type worms (including the one isolated from a human host) in the heterozygosity analysis we noticed that they showed very high apparent heterozygosity. Strikingly, this was also the case for heterozygosity on the X chromosome in the male worm. To determine if this was an anomaly of our sample from Bangladesh we subjected the sequences of the five whole genome sequenced "dog only" type worms from (Jaleta et al., 2017) to the same analysis (Figure 8 in the body of the publication and Fig. Suppl_File_3_1). Except for the worm L6, these sequences showed even higher apparent heterozygosity, including on the X chromosome (all "dog only" type whole genome sequenced individuals by (Jaleta et al., 2017) were males). Notice that with respect to the mitochondrial and the nuclear genomes, L6 was the only whole genome sequenced worm in the study of (Jaleta et al., 2017) that belongs to a separate sub-cluster of the "dog-only" cluster, of which it is not clear, if it is more closely related to the other "dog only" cluster (to which the other four worms and the worms isolated in this study belong) or to the "human and dog" cluster (compare the positions of L6 in Figures 4-6 in the main body of the paper and Suppl. Figures 1, 2). For the rest of this Suppl. File 3 "dog only" always means "dog only" except for L6. L6 is not included in this analysis.

First, we asked if some of the *S. stercoralis* in Asia, in particular the "dog only" type might not employ XX/XO sex determination as it is the case in the USA derived *S. stercoralis* reference isolate (Hunt et al., 2016). We therefore performed read coverage analysis and compared "dog only" cluster males, which have the high apparent heterozygosity on the autosomes and the X chromosome, with males and females from the reference isolate (which have low heterozygosity) and from "human and dog" cluster sequences from China, which have high heterozygosity but do not show the unexpected apparent heterozygosity on the X chromosome (Fig. Suppl_File_3_2). The proportional X-chromosome coverage in all males was comparable and clearly less than in females, suggesting that all these males do have only one X chromosome. Notice that free-living adults contain highly polyploid nuclei in the distal part of their gonads, in which the X chromosome is less amplified, compared with the autosomes in both sexes (Hammond and Robinson, 1994; Kulkarni et al., 2016). Therefore, the overall coverage of the X chromosome in free-living adult females is less than 100% (compared with autosomes) but still more than 50% (in our cases between 70 and 80%) and less than the expected 50% in males (Hunt et al., 2016; Kulkarni et al., 2016). In infective larvae, all of which are female and do not have the polyploid nuclei yet, the coverage of the X is equal to the autosomes (Fig. Suppl_File_3_2E)

We then analysed the heterozygosity over the length of the chromosomes. Males did indeed show low heterozygosity over large portions of the X chromosome compared with females and with autosomes of both sexes (Fig. Suppl_File_3_3). However, there are apparent heterozygosity hot spots that dominated the analysis. These hotspots are also present in females. Also on autosomes, such heterozygosity hotspots are visible, although less frequent. We think these hotspots reflect duplications and X to autosome translocations present in the genome of the "dog only" type, compared with the *S. stercoralis* reference genome (Hunt et al., 2016).

Overall, we think that the heterozygosity detected in the "dog only" worms is at least partially artificial due to the use of a too different reference genome that does not contain some duplications and X to autosome translocations present in these worms. Currently we cannot really tell what portion of the apparent heterozygosity is real and how much is artificial. The problem appears to be aggravated on the X chromosome because also females show higher heterozygosity on the X chromosome than on the autosomes. Also, in the reference genome, the assembly of the autosomes is better than the one of the X chromosome. Some of the low but detectable apparent heterozygosity seen on the X in males of the human derived *S. stercoralis* might also be caused artificially due to assembly issues.

**Figure legends**

Fig. Suppl_File_3_1: Apparent heterozygosity of "human and dog" type worms from this and previous studies [(Aupalee et al., 2020) Thailand, (Kikuchi et al., 2016) Japan and Myanmar,  (Beiromvand et al., 2024) Iran, (Zhou et al., 2019) China, (Jaleta et al., 2017) Cambodia and (Hunt et al., 2016) USA] plus the "dog only" type worms from this study and from Jaleta et al. (2017). The X axis shows the heterozygosity on the autosomes, the Y axis the heterozygosity on the X chromosome. The diagonal indicates equal heterozygosity on the X chromosome and the autosomes. The labels only refer the circled worms in the same colour as the label. Notice the high heterozygosity on the X chromosome in males (squares) of the "dog only" type. In particular, compare the human derived worms with hight heterozygosity from China (green) with the dog derived samples.  For a discussion of the one female in the male group from China see Zhou et al. (2019). Notice that for the "dog only" worms the females are above the diagonal while the males are on or below the diagonal, indicating inflated X chromosomal heterozygosity in females as well. For the details of the analysis see Materials and Methods in the main text of this publication.

Fig. Suppl_File_3_2: Read coverage of individual male and female worms. The reads from single worm Illumina sequencing were aligned with the reference genome (Hunt et al., 2016) and for each position the coverage was determined. The x-axes show the coverage, the y-axes show the number of positions with the corresponding coverage for the autosomes (blue) and the X chromosome (red). the % indicate the relative coverage of the X chromosome compared with the autosomes. Notice that due to the uneven amplification of the genomic DNA in the highly polyploid nuclei of the distal gonad the X chromosome is underrepresented in free living adults of both sexes  (Hunt et al., 2016; Kulkarni et al., 2016). (A,B) females  and (C,D) males of the reference isolate (data from Hunt et al. (2016). (E-H) worms from the study by Zhou et al. (2019) in China. (E) infective larva, which is female but has no polyploid germline cells. (F-H) males that have high heterozygosity on the autosomes but not on the X chromosome. The coverage of the X chromosome is comparable with the one in the reference males, confirming that these males have only one X chromosome. Notice that no free-living females were found by Zhou et al. (2019). (I,K) dog only type worms from Jaleta et al., (2017) that have high apparent heterozygosity on the autosomes and on the X chromosome (Fig. Suppl_File_3_1). The coverage of the X chromosome is comparable with the reference males suggesting these males have only one X chromosome. Notice the rather high number of low or completely not covered positions and the irregular shape of the graph towards the left of the graph. We think that this is caused by the rather large sequence and genome structural differences between these sequences and the reference sequence. The sequencing depth in the Bangladesh study (this manuscript) was lower such that a reliable quantitative comparison between the sexes was not possible, but the coverage of the X in males was clearly lower than in females in this study as well.

Fig. Suppl_File_3_3: Distribution of heterozygous positions over the three largest X chromosomal contigs (A) and over the two largest autosomal scaffolds (B). The x-axes show the position along the contig/scaffold and the y-axes show the number of apparently heterozygous positions per 10 kb window. The black lines represent the running means. Three females and the one male from the "dog only clade" of this study are shown. Notice the very low number of heterozygous positions on the X chromosome in the male over most of the X chromosomal contigs and the apparent hotspots (arrows). The same hotspots are also visible in the females. Hotspots are also present on the autosomes, although less frequent (notice that the autosomal scaffolds are much longer than the X chromosomal ones.

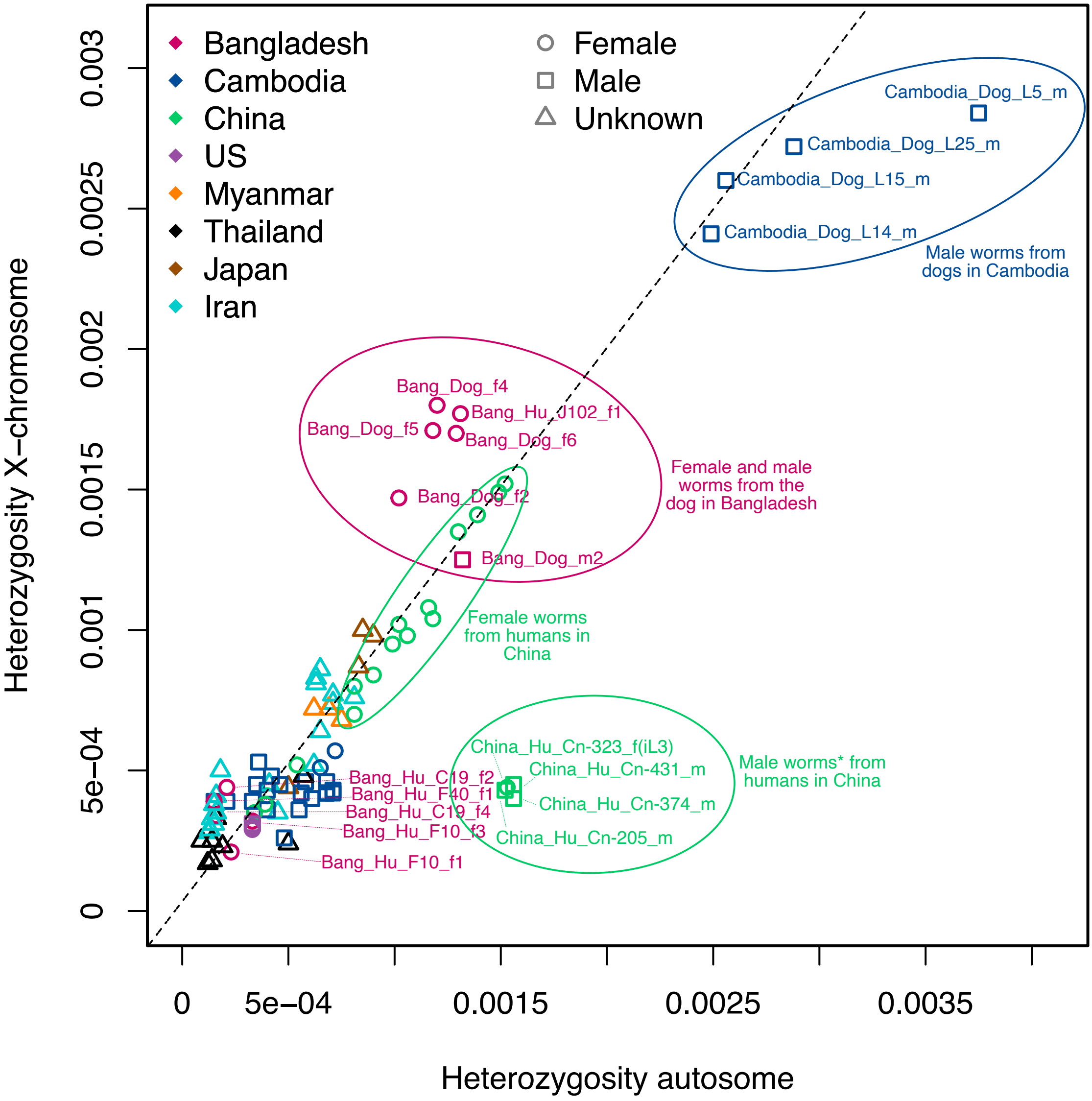
Bangladesh
Cambodia
China
US
Myanmar
Thailand
Japan
Iran
Female
Male
Unknown
Cambodia_Dog_L5_m
Cambodia_Dog_L25_m
Cambodia_Dog_L15_m
Cambodia_Dog_L14_m
Male worms from dogs in Cambodia
Bang_Dog_f4
Bang_Hu_J102_f1
Bang_Dog_f5
Bang_Dog_f6
Bang_Dog_f2
Bang_Dog_m2
Female and male worms from the dog in Bangladesh
Female worms from humans in China
China_Hu_Cn-323_f(iL3)
China_Hu_Cn-431_m
China_Hu_Cn-374_m
China_Hu_Cn-205_m
Male worms* from humans in China
Bang_Hu_C19_f2
Bang_Hu_F40_f1
Bang_Hu_C19_f4
Bang_Hu_F10_f3
Bang_Hu_F10_f1
Heterozygosity X−chromosome
Heterozygosity autosome
0
5e−04
0.001
0.0015
0.002
0.0025
0.003
0.0035

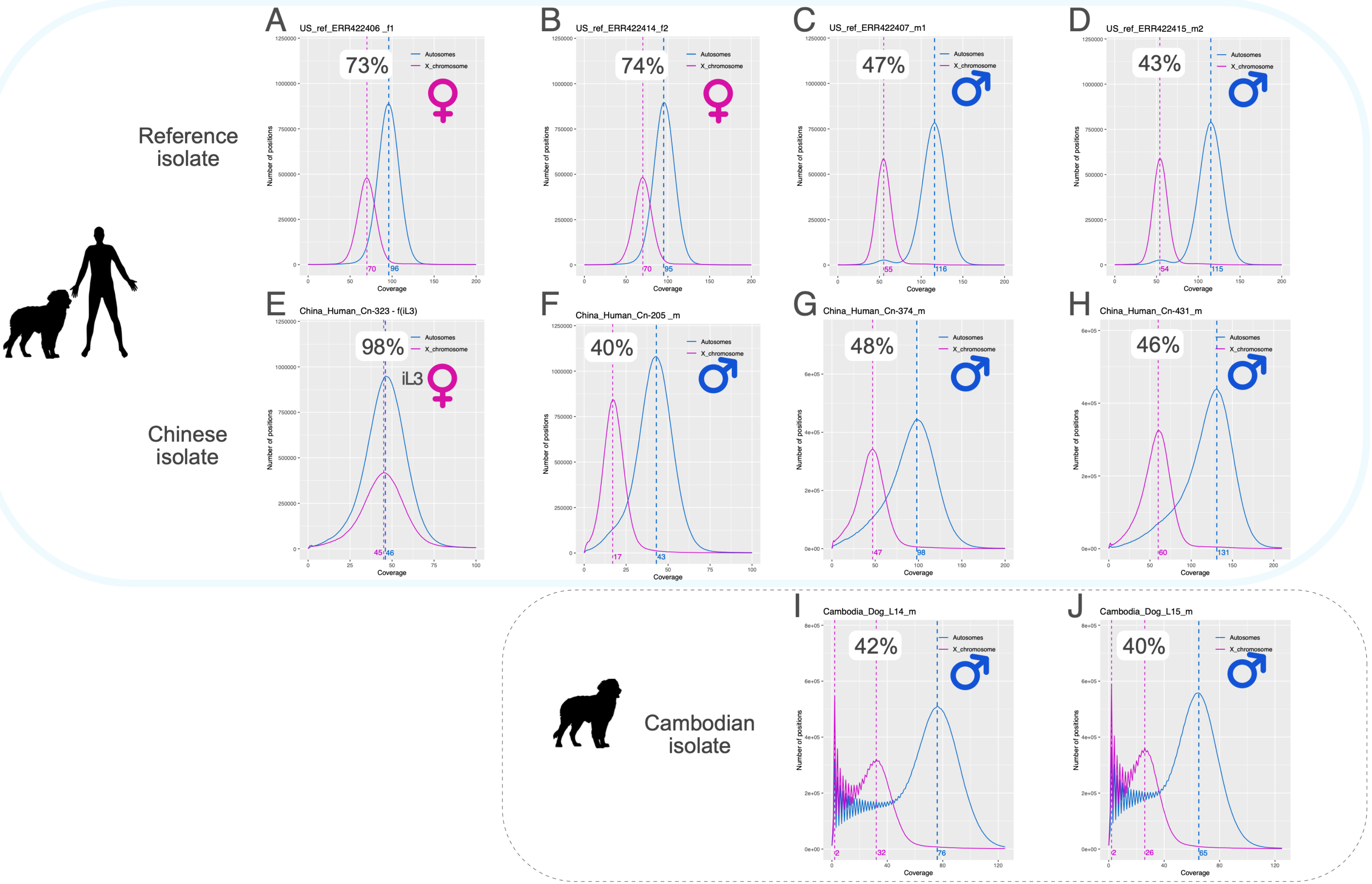

Reference isolate
Chinese isolate
Cambodian isolate
A US_ref_ERR422406 _f1
73%
B US_ref_ERR422414_f2
74%
C US_ref_ERR422407_m1
47%
D US_ref_ERR422415_m2
43%
E China_Human_Cn-323 - f(iL3)
98%
iL3
F China_Human_Cn-205 _m
40%
G China_Human_Cn-374_m
48%
H China_Human_Cn-431_m
46%
I Cambodia_Dog_L14_m
42%
J Cambodia_Dog_L15_m
40%
Autosomes
X_chromosome
Number of positions
Coverage

Fig_Suppl_file_3 3

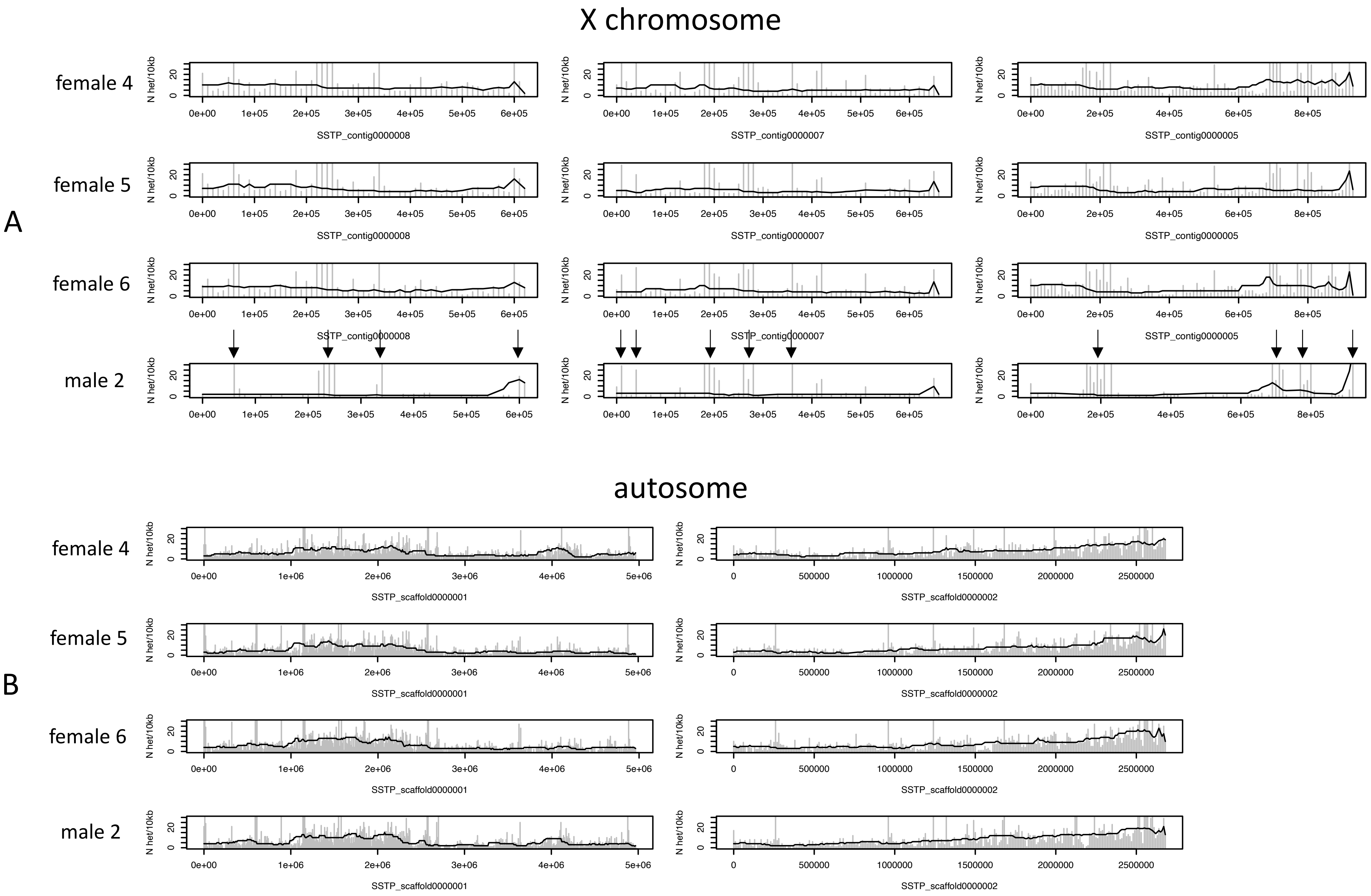
